# When Can Elimination of SARS-CoV-2 Infection be Assumed? Simulation Modelling in a Case Study Island Nation

**DOI:** 10.1101/2020.05.16.20104240

**Authors:** Nick Wilson, Matthew Parry, Ayesha J Verrall, Michael G Baker, Markus Schwehm, Martin Eichner

## Abstract

**Aims:** We aimed to determine the length of time from the last detected case of SARS-CoV-2 infection before elimination can be assumed at a country level in an island nation.

**Methods:** A stochastic version of the SEIR model Covid SIM v1.1 designed specifically for COVID-19 was utilised. It was populated with data for the case study island nation of New Zealand (NZ) along with relevant parameters sourced from the NZ and international literature. This included a testing level for symptomatic cases of 7,800 tests per million people per week.

**Results:** It was estimated to take between 27 and 33 days of no new detected cases for there to be a 95% probability of epidemic extinction. This was for effective reproduction numbers (Re) in the range of 0.50 to 1.0, which encompass such controls as case isolation (the shorter durations relate to low Re values). For a 99% probability of epidemic extinction, the equivalent time period was 37 to 44 days. In scenarios with lower levels of symptomatic cases seeking medical attention and lower levels of testing, the time period was up to 53 to 91 days (95% level).

**Conclusions:** In the context of a high level of testing, a period of around one month of no new notified cases of COVID-19 would give 95% certainty that elimination of SARS-CoV-2 transmission had been achieved.

## Introduction

Some nations are attempting to eliminate the SARS-CoV-2 pandemic virus as in the case of New Zealand^1^ and for Australia this has been articulated as a potential option.^2^ It might also be the goal for the island jurisdiction of Taiwan and for South Korea, which has well controlled land borders (though neither of these places appears to have actually specified this goal).

Once a nation can declare itself having achieved elimination status it can potentially phase out restrictive disease control measures within the country, while maintaining tight border controls with quarantine for incoming travellers. It could also permit quarantine-free travel with other COVID-19 free nations, as envisaged by the Prime Ministers of Australia and New Zealand in terms of a trans-Tasman “bubble”.^3^ Similarly, the leaders of Austria, Greece, Israel, Norway, Denmark, the Czech Republic, Singapore, Australia and New Zealand “agreed that as each begins to ease restrictions they could capitalise on low infection rates by creating tourism safe zones”.^4^

We selected New Zealand as a case study island nation as it has an elimination goal,^1^ has good border controls and is making steady progress towards elimination with occasional days in May 2020 with no new cases reported.^5^ Some work has already been done on the elimination topic in New Zealand, with a modelling group^6^ reporting that a “90 day period at Level 4 [intensive lock-down restrictions] leads to containment to very low levels by the end of the 90 day period”. “This scenario leads to elimination in approximately 50% of stochastic realisations”. Another study^7^ estimated the probability that regions within New Zealand might have achieved elimination, with the District Health Board region with the highest number of days free of notified cases (Wairarapa at 16 days as of 18 April 2020) having a 92% probability of having eliminated the pandemic virus. However, none of this work has defined time periods for differing levels of probability for elimination being achieved at a whole country level. Estimating these periods was therefore the aim of this simulation study.

## Methods

To run “time to elimination” analyses for New Zealand, we used a stochastic SEIR type model with key compartments for: susceptible [S], exposed [E], infected [I], and recovered/removed [R]. The model is a stochastic version of CovidSIM which was developed specifically for COVID-19 (http://covidsim.eu; version 1.1). Work has been published from previous versions of this model,^8 9^ but in the Appendix we provide updated parameters and differential equations for version 1.1. A similar simulation approach was taken previously for a poliomyelitis elimination study.^10^ The stochastic model was built in Pascal and 10,000 simulations were run for each set of parameter values.

The parameters were based on available publications and best estimates used in the published modelling work on COVID-19 (as known to us on 13 May 2020). Key components were: a starting position of 10 infected cases, the assumption of effective border control with no new imported cases, 80% of infected COVID-19 cases being symptomatic, 39.5% of cases seeking medical consultation in primary care settings, and 4% of symptomatic cases being hospitalised (see Table A1 in the Appendix for the full set of parameters used). The level of testing of symptomatic cases in primary care and for respiratory cases being tested when hospitalised (both at 95% coverage with test sensitivity of 89%^11^), was as per a fairly optimal surveillance system conducting 7,800 tests per million people per week (slightly less than the level in New Zealand as per mid-May 2020 at 8,190 tests per million people per week^12^). Identified cases were transferred to supervised isolation which was assumed to be 95% effective. We considered different levels of transmission with the effectivere production number (Re) of SARS-CoV-2 to be 0.5, 0.6, 0.7, 0.8, 0.9, and 1.0 (with extinction still occurring in the last case [Re = 1.0] due to accumulation of immune individuals after infection). These levels of Re were assumed to represent the summated total of pandemic-related physical distancing (voluntary and mandated), travel restrictions, hygiene behaviours, mask use, voluntary staying at home when unwell, and contact tracing resulting in quarantine of contacts.

In each stochastic simulation, the delay since the last detected cases was stored on a daily basis together with the information as to whether infection was still ongoing or was extinguished. The probability that extinction had occurred if no cases have been reported for a given number of days was calculated by dividing the number of such delays during extinction by the total number of such delays (ie, with ongoing transmission or with extinction). Other scenarios considered the impact of higher starting numbers of infected cases, and lower levels of attendance for medical consultations in primary care and also for the level of testing.

## Results

It was estimated to take between 27 and 33 days of no new detected cases for there to be 95% probability of epidemic extinction for effective reproduction numbers (Re) in the range of 0.5 to 1.0, combined with effective case isolation (Table 1; the shorter durations relate to low Re values as depicted in Figure 1). This range was 37 to 44 days (around 5 to 6 weeks) for the 99% probability level. In scenarios with lower levels of symptomatic cases seeking medical attention and lower levels of testing, the time period was up to 53 to 91 days (second scenario in Table 1; Figure 2; for the 95% level of probability). Starting the simulations with 100 or 1000 initially infected cases instead of 10, made no meaningful difference (Figure 2). This was expected because while there are 1000 or 100 infected individuals in the population, the frequency of very short case-free intervals of up to a few days at most are very frequent whereas long case free intervals of weeks are virtually impossible. Only after the number of cases has dropped to very low numbers, case-free periods of weeks may occasionally occur despite ongoing transmission. Also only for such long case-free periods is there an ambiguity whether extinction has occurred or not, whereas nobody would assume that extinction has occurred if no cases have been reported for two or three days.

**Table 1:** Probability level of SARS-CoV-2 elimination by number of days since the last detected case of COVID-19 (starting with 10 infectious cases for each value of Re in the range of: 0.5 to 1.0; using 10,000 simulations per parameter set).

| Base case and scenarios | 50% probability | 95% probability |
| --- | --- | --- |
| Base case analysis: 39.5% of symptomatic cases seek medical attention in primary care [Flutracking data for NZ] where 95% of them are tested | 12 to 15 days | 27 to 33 days |
| Scenario 1: 30% seek attention, 60% of them are tested | 17 to 24 days | 38 to 51 days |
| Scenario 2: 20% seek attention, 30% of them are tested | 24 to 42 days | 53 to 91 days |

**Figure 1:**
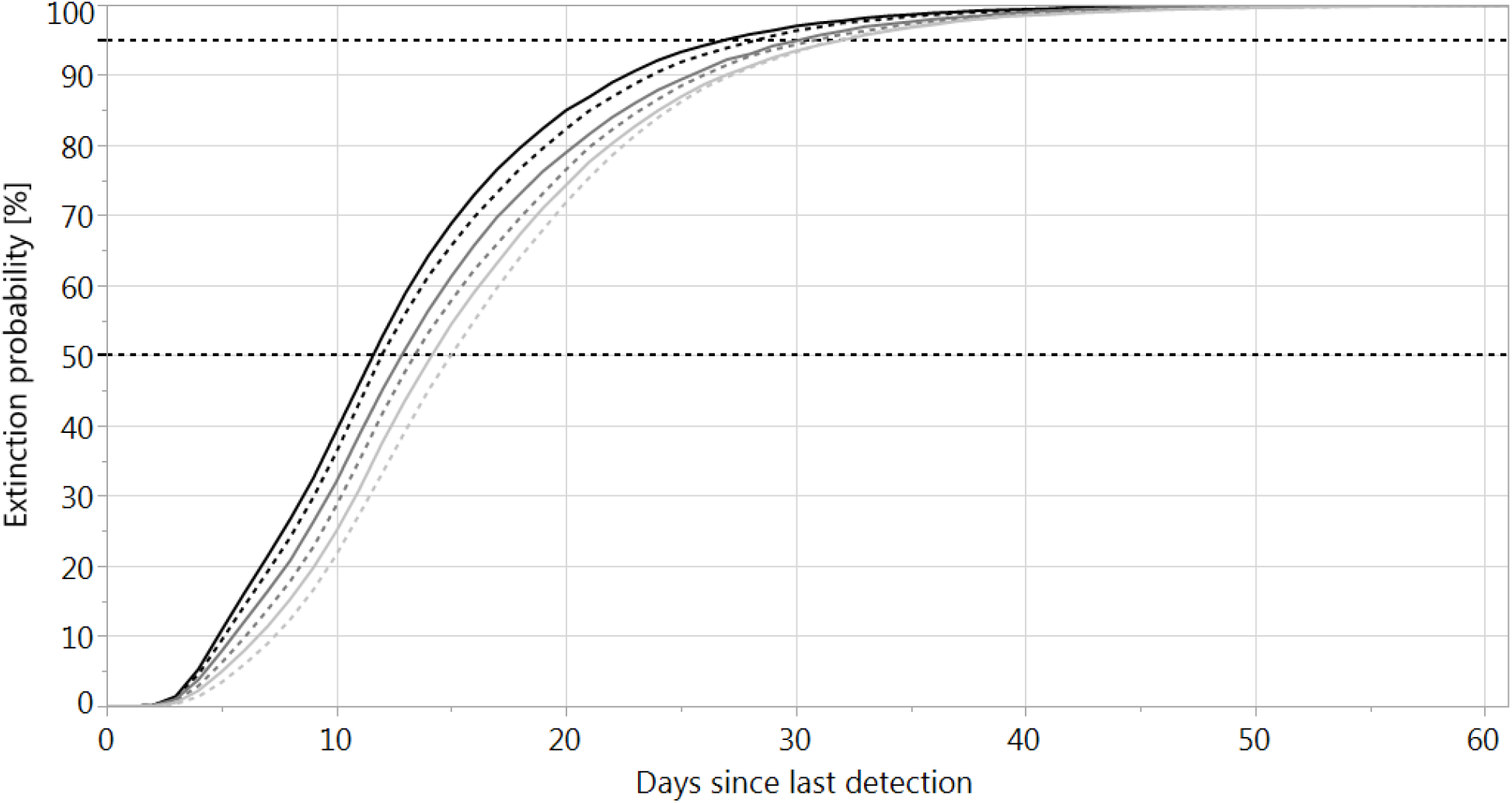
Probability that pandemic virus extinction has occurred according to different Re values. The chart shows extinction probabilities if no new COVID-19 cases have been reported for a given number of days (10,000 simulations starting with 10 infectious cases for each value of Re). The curves from left to right relate to Re = 0.5, 0.6, 0.7, 0.8, 0.9, 1.0, respectively.

**Figure 2:**
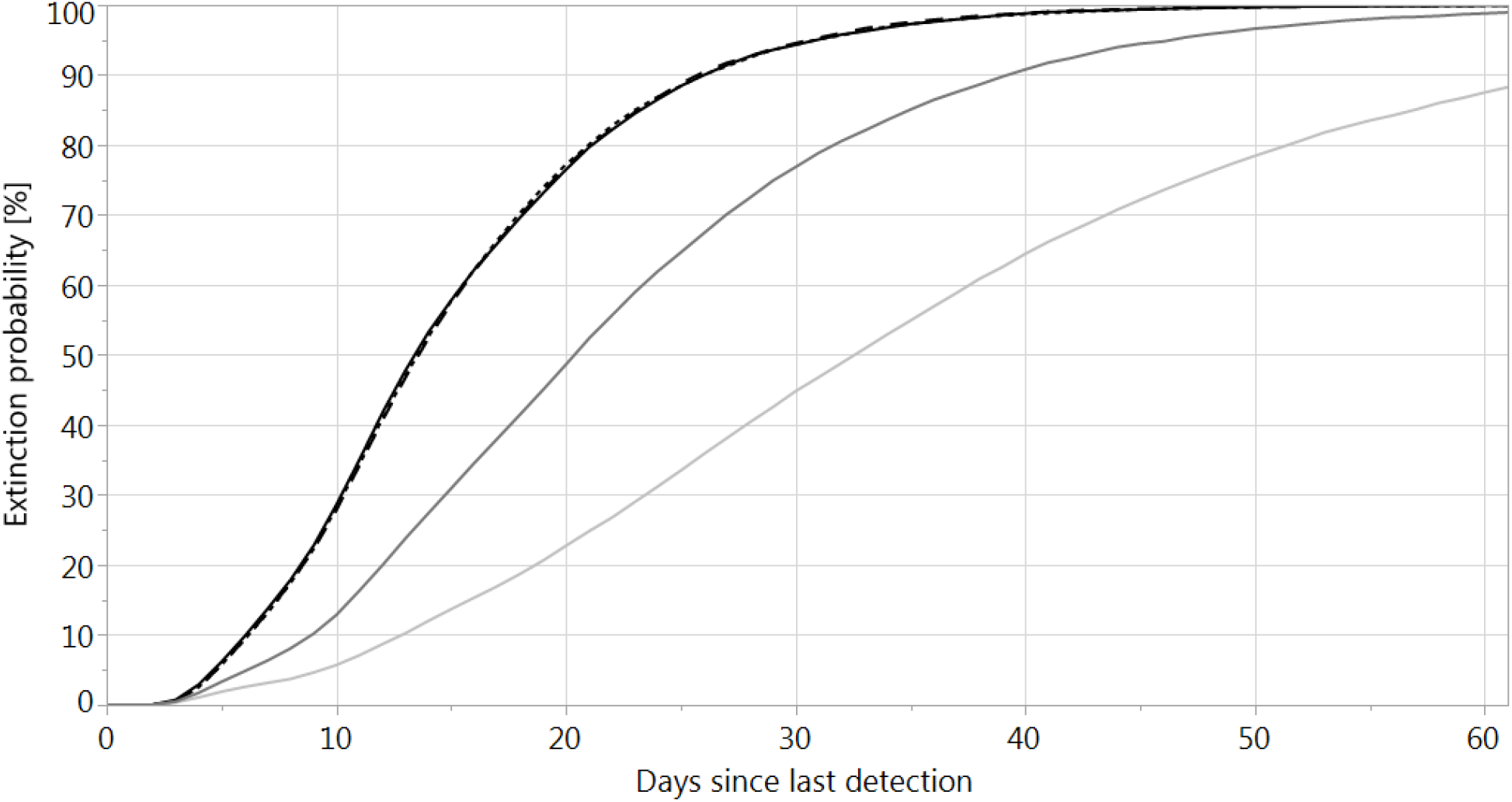
Probability that pandemic virus extinction has occurred according to different numbers of initial cases and testing proportions. The chart shows extinction probabilities if no new COVID-19 cases have been detected for a given number of days (effective reproduction number Re=0.8; 10,000 simulations each scenario). The three black curves on the left (nearly undistinguishable) give the results obtained when starting the simulations with 10 (solid), 100 (dotted) or 1,000 (dashed) infections, respectively. The dark grey curve shows the result when only 30% of COVID-19 patients seek medical attention and only 60% of them are tested (Scenario 1). The light grey curve on the right shows the result when only 20% seek medical attention and only 30% of them are tested (Scenario 2).

## Discussion

These simulations suggest that with an effective surveillance system (high levels of testing symptomatic cases with cough and fever and testing of respiratory cases being hospitalised), it is possible to develop high assurance that an elimination goal has been achieved in around four to five weeks of no cases being detected (Figure 1). Of course this modelling assumes that Re values are being successfully suppressed (at Re ≤ 1.0), which may be feasible if there are both rigorous public health control measures (contact tracing, case isolation etc) and people maintain some levels of behaviour change in the peri-elimination period (hand and respiratory hygiene, mask use, working from home where feasible, staying home when unwell etc).

The time period for no new cases we have identified of four to five weeks is similar to a provisional “28 days since the onset date of the last known infection” that has been suggested for New Zealand previously.^13^ This is also the span of two maximal incubation periods (14 days, which is being used for determining the length of quarantine for incoming travellers in New Zealand). A 28 day period for elimination has also been proposed for the Australian setting.^2^ Nevertheless, it is possible that the 99% level of probability provides a better safety margin (ie, the 37 to 44 day period we have estimated). This would be more appropriate if there are uncertainties with the distribution of testing by geographical region and socio-demographic group. It might also be considered more appropriate by policy makers wanting to reduce international travel restrictions between COVID-19 free nations (see *Introduction*).

A limitation of our work is that we didn’t perform scenario analyses that captured differing performance of the contact tracing system – instead we assumed that effective routine contact tracing was part of the low Re values. A more sophisticated model (eg, an agent based SEIR model) would be needed to separate out the effect of contact tracing. Another limitation was that we didn’t consider extreme scenarios such as residual transmission amongst groups of school children where infection is more likely to be asymptomatic or only mildly symptomatic (ie, this could result in further delays in detection).

## Conclusion

In the context of a high level of testing, a period of around one month of no new notified cases of COVID-19 would give 95% certainty that elimination of SARS-CoV-2 transmission had been achieved.

## Data Availability

All the Excel files with the results are available from the authors on request

## Acknowledgements

Dr Schwehm is supported by the University of Tübingen and the IMAAC-NEXT Association. Professor Wilson is supported by the New Zealand Health Research Council and Ministry of Business Innovation and Employment (MBIE) funding of the BODE3 Programme.

## Appendix: Mathematical description of the CovidSIM model (version 1.1) and model parameters

The stochastic simulations are based on the following differential equations:

### Model dynamics

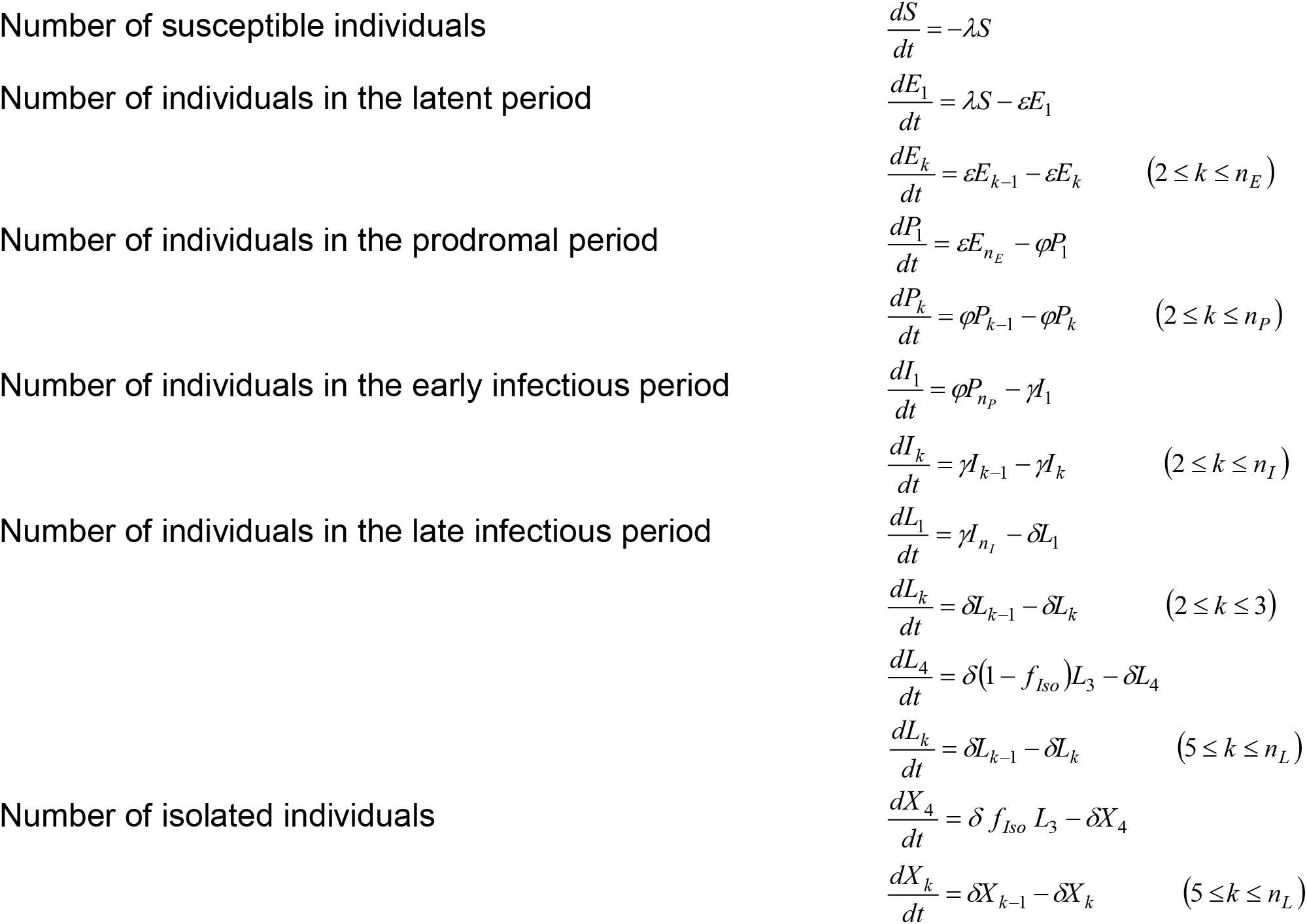

### Derived variables

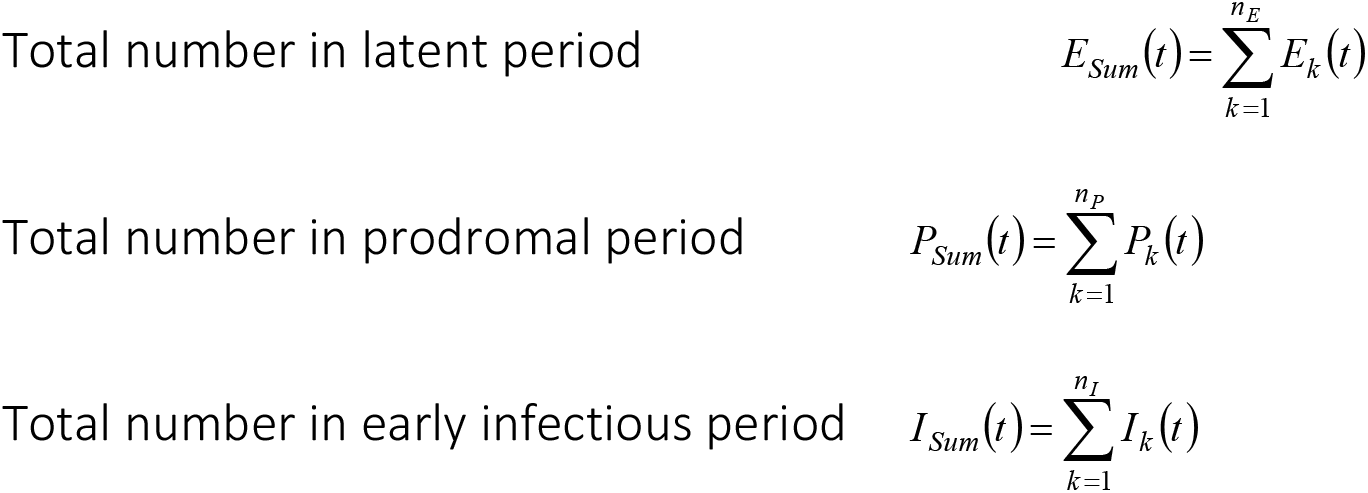
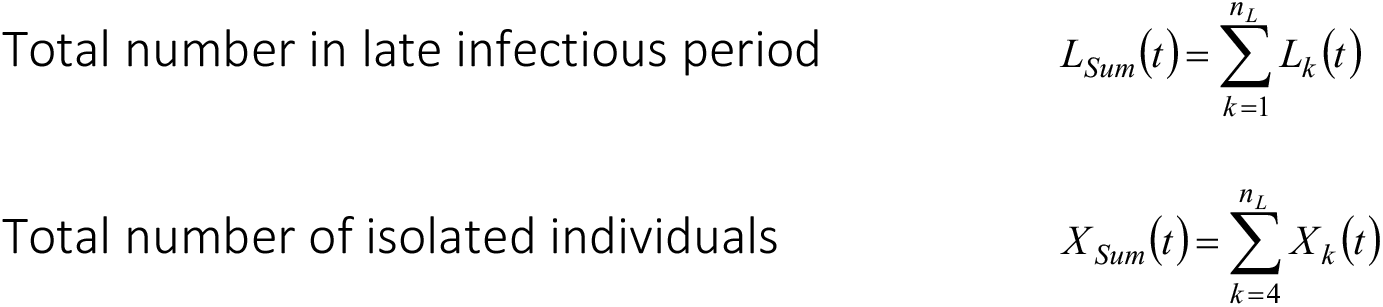

### Contact rate and force of infection

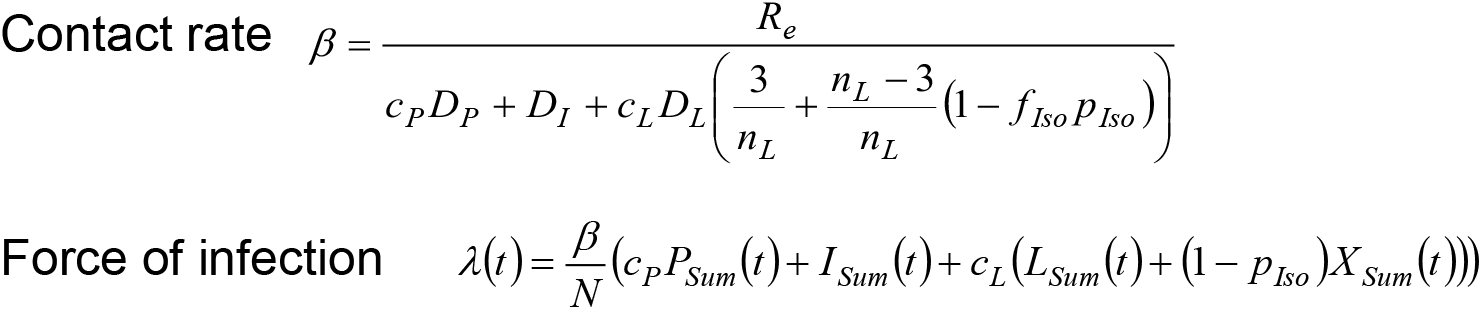

### Stochastic treatment of the differential equations

The kind of epidemiologic events and the duration between two consecutive events are calculated using random numbers. The simulations start with a susceptible population with a given number of infected individuals (“index cases”). The individual infection stages of these index cases are picked at random, taking into consideration the different lengths of the latent, prodromal, early and late infectious period. In each simulation, the sum of all the rates that change the current state of the system is calculated as

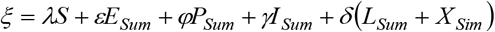

A uniformly distributed random number *r*_1_ ∈[0, *ξ*] is then chosen, and the time Δ*t* = −ln(*r*_1_)/*ξ* after which the next event occurs is calculated. All transition rates are arranged in an arbitrary order, and cumulative rates are calculated by adding their individual rates. A new uniformly distributed random number *r*_2_ ∈[0, *ξ*] is chosen, and the first transition in the order whose cumulative rate is larger than *r*_2_ is performed. If, for example, the event is an infection, one individual is removed from the group of susceptible individuals and added to the group of latent individuals of stage 1. If a transition is scheduled to take place to an individual in stage *L*_3_, a third random number is calculated to determine whether the case is diagnosed and isolated or whether the case progresses to *L*_4_. New rates are calculated after each step, and the procedure is repeated. A more detailed description of the transformation of differential equation models to stochastic models can be found in Gillespie (1976).^14^

### Parameters

*N* Population size
*λ* Force of infection
*R_e_* Effective reproduction number
*β* Effective contact rate
*D_E_* Average duration of the latent period
*n_E_* Number of stages for the latent period
*ε* Stage transition rate for the latent period (*ε* = *n_E_/D_E_)*
*D_P_* Average duration of the early prodromal period
*n_P_* Number of stages for the early prodromal period
*φ* Stage transition rate for the early prodromal period (*φ* = *n_P_* / *D_P_)*
*c_P_* Contagiousness in the prodromal period (relative to the contagiousness in the early infectious period)
*D_I_* Average duration of the early infectious period
*n_I_* Number of stages for the early infectious period
*γ* Stage transition rate for the early infectious period (*γ* = *n_I_* / *D_I_*)
*D_L_* Average duration of the late infectious period
*n_L_* Number of stages for the late infectious period
*δ* Stage transition rate for the late infectious period (*δ* = *n_L_* / *D_L_*)
*c_L_* Contagiousness in the late infectious period (relative to the contagiousness in the early infectious period)
*f_Sick_* Fraction of individuals in the (early and late) “infectious period” who have symptoms (i.e. who are sick)
*f_Consult_* Fraction of sick cases who seek medical help
*f_Hosp_* Fraction of sick cases who are hospitalized
*f_Test_* Tested fraction of cases who seek medical help or who are hospitalized
*s_Test_* Sensitivity of the test
*f_Iso_* = *f_Sick_*(*f_Consult_+f_Hosp_*)*f_Test_s_Test_* Fraction of infected individuals who are isolated due to a positive test result
*p_Iso_* Fraction of contacts which are prevented for isolated cases

**Table A1:**
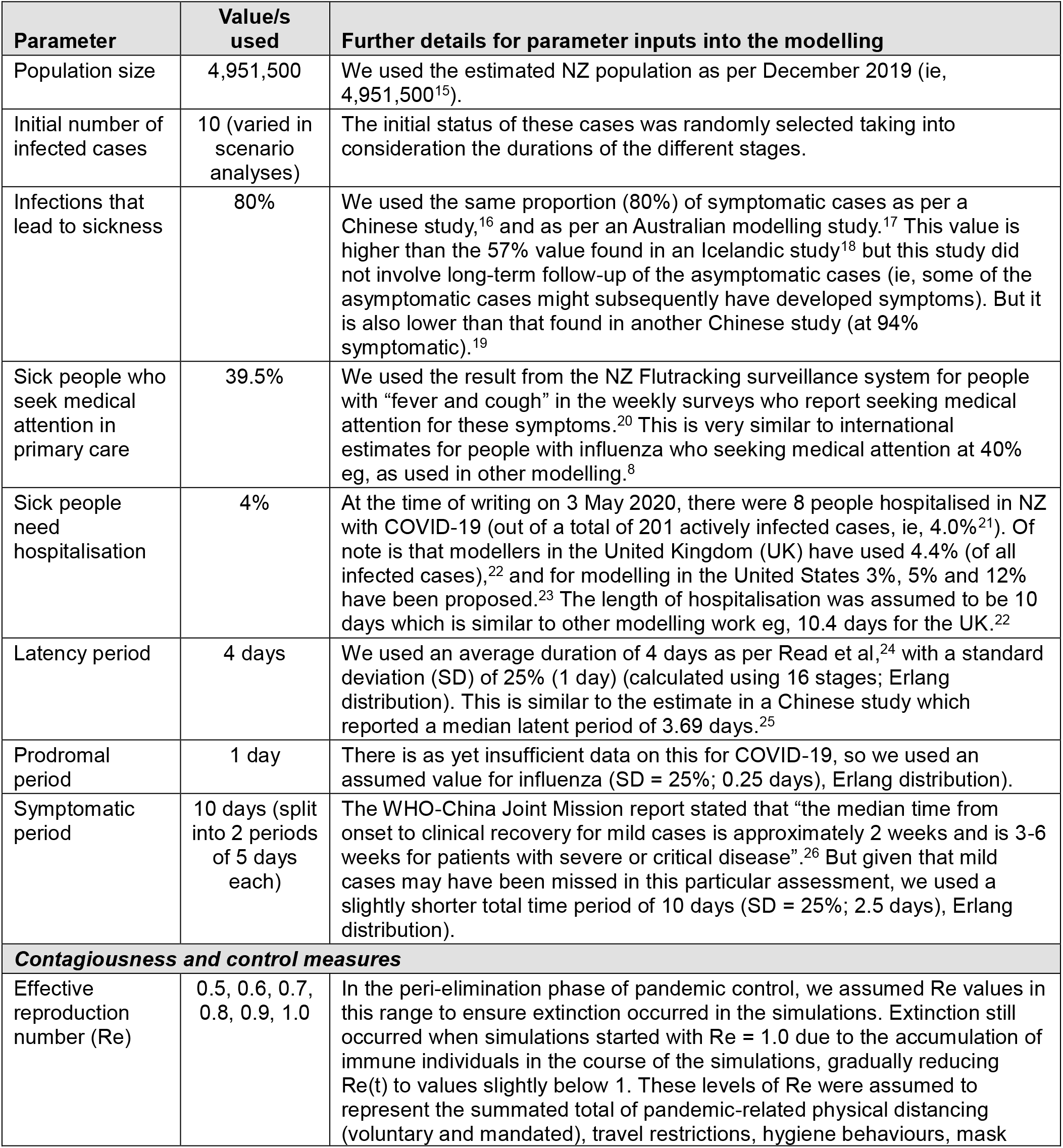
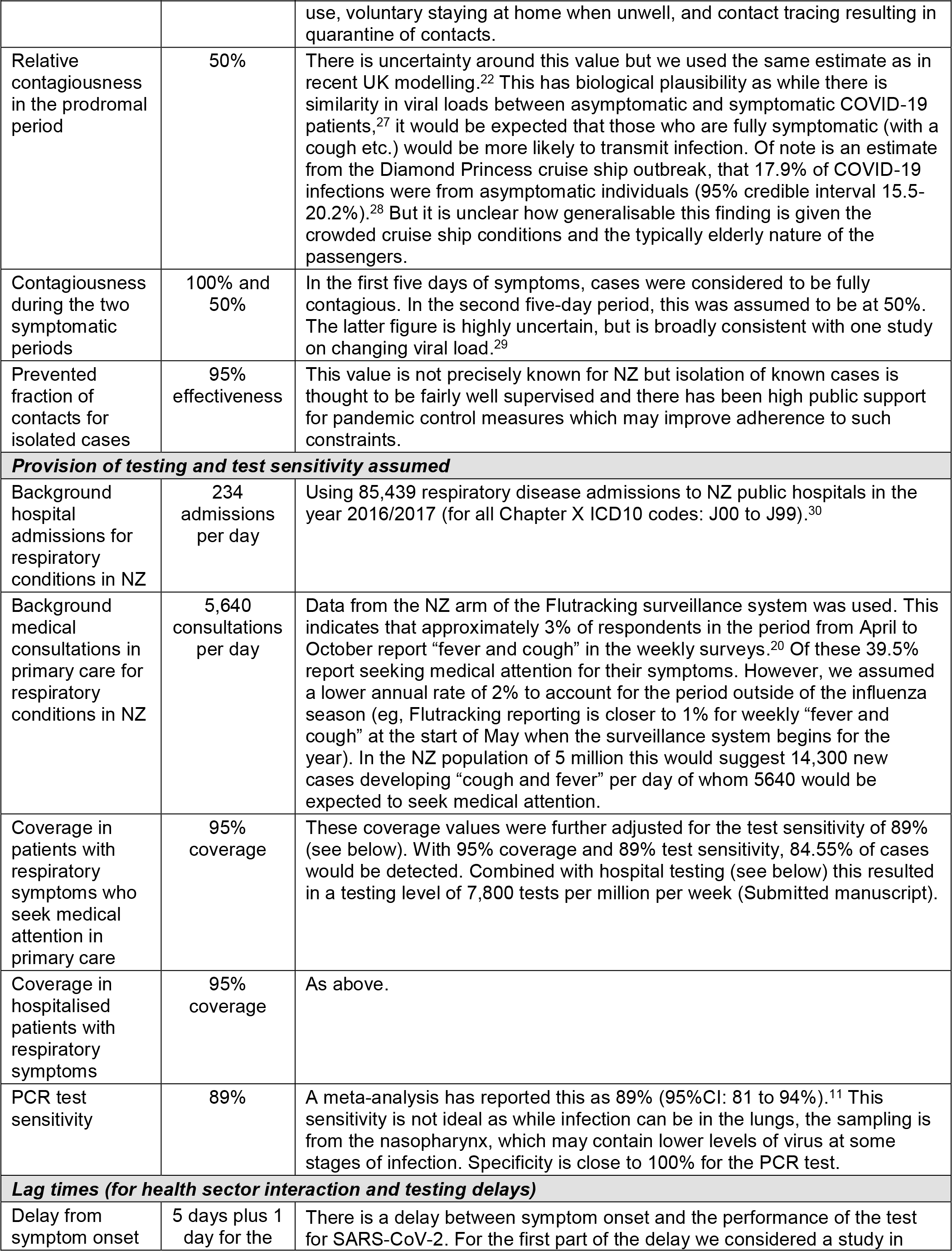
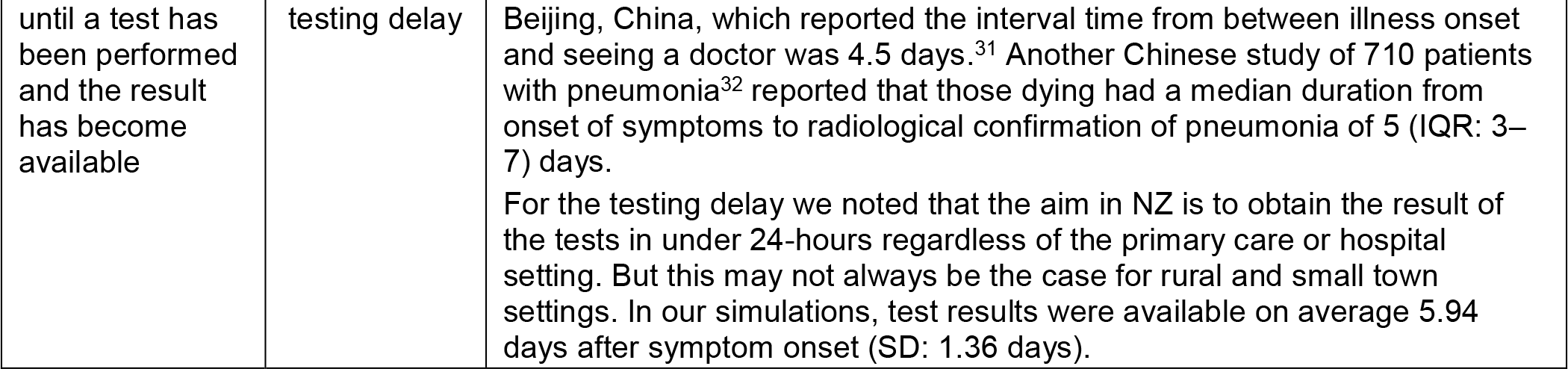
Input parameters used for modelling the potential spread of the COVID-19 pandemic with the stochastic version of CovidSIM (v1.1) with New Zealand as a case study.

